# Personal-MetaboHealth, an actionable health check in middle age, is improved by an effective lifestyle intervention in those at risk

**DOI:** 10.64898/2026.02.15.26346369

**Authors:** Niels van den Berg, Gabriela Natalle Lopes, Fatih Bogaards, Marian Beekman, Edson Amaro Junior, Joris Deelen, P. Eline Slagboom

**Author notes:** Corresponding authors, Niels van den Berg;, Joris Deelen. These authors contributed equally (Niels van den Berg, Gabriela Natalle Lopes). These authors contributed equally (Joris Deelen, P. Eline Slagboom).

## Abstract

The biomarker MetaboHealth represents a novel indicator of overall health in middle age and may potentially be suitable as actionable health check in prevention strategies. MetaboHealth is a blood-based metabolomic composite score that predicts a wide range of age-related conditions and mortality in large European cohorts. Here, we investigated whether MetaboHealth can be personalised and limited to clinically validated metabolomic markers. Next, we assessed whether the updated MetaboHealth score predicts all-cause mortality and cardiometabolic disease incidence and can be improved by a lifestyle intervention. To personalise MetaboHealth, we scaled the metabolomic markers using a Dutch reference population (i.e. the Biobanking and BioMolecular Research Infrastructure Netherlands) and, in addition, based the score solely on clinically validated metabolic markers. The novel version of the score, Personal-MetaboHealth, retained predictive accuracy for all-cause mortality and showed an even stronger association with incident cardiometabolic disease in the Leiden Longevity Study (LLS) in which 2,404 participants were followed for up to 22 and 16 years for mortality and morbidity, respectively. The association of Personal-MetaboHealth with all-cause mortality remained robust after adjusting for smoking, alcohol use, and medication, while the cardiometabolic disease association was partially driven by smoking. Each standard deviation decrease in Personal-MetaboHealth was associated with a 11.7 year earlier onset of the first cardiometabolic disease in the LLS. Next we showed that Personal-MetaboHealth can be improved by a 3-month combined lifestyle intervention in middle aged individuals (Growing Old Together study), specifically in those at risk with an unhealthy score at baseline. Personal-MetaboHealth thus offers a potential actionable health check in middle age for early prevention and extension of healthy lifespan.

## Introduction

The global population is undergoing a demographic shift towards a high proportion of older age groups, such that nearly one in five persons will be aged 60 years or older by 2030 (https://platform.who.int/data/maternal-newborn-child-adolescent-ageing/indicator-explorer-new/mca/percentage-of-total-population-aged-60-years-or-over). In parallel to this demographic shift, our lifespan continues to increase while gains in years lived free of disease have been more limited, resulting in prolonged morbidity and disability. The co-occurrence of this demographic shift and slower gains in disease-free life years places a significant burden on healthcare systems and results in high costs[1], underscoring the importance to improve quality of life and extend the years lived in good health[2]. Therefore, preventive strategies must move beyond symptom-based detection and instead focus on the early identification of individuals at increased risk of age-related conditions (e.g. decline of cognitive, metabolic, and muscle function), diseases, and multi-morbidity, while clinical symptoms have not manifested yet. Markers allowing such early identification are necessary in order to professionally support evidence-based lifestyle programs.

Besides a diagnosis as wake-up call for a lifestyle change, affordable, scalable, and validated molecular markers may serve as motivational tools for those at risk in the prevention of overall health decline in the population^3^. Recent technological advances, such as standardised -omics-based measurements, offer novel opportunities to generate biomarkers that capture age-related systemic changes that precede disease onset, opening a window of opportunity for prevention[3]. Although many overall health scores, aiming to capture ageing-related decline, have been constructed, most are based on cross-sectional data or are expensive to generate[3]. For this reason, the MetaboHealth score was developed as a cost-efficient and scalable high-throughput nuclear magnetic resonance (¹H-NMR) spectroscopy measurement-based biomarker indicating overall health. MetaboHealth was trained on prospective mortality data from 12 large cohorts and was generated to predict mortality risk in population-based cohorts[4], which it does at all ages, but particularly successful in the middle-aged population. Beyond accurately predicting mortality risk, the MetaboHealth score is associated with a wide range of age-related conditions and functional decline, including somatic frailty, COVID-19-related outcomes, decreased cognitive processing, poorer memory, and reduced functional independence[5–8]. Together, these findings established MetaboHealth as a valuable tool for assessing population-level differences in overall health and a potential health check for prevention through lifestyle programs.

Applications of MetaboHealth, as well as other overall health scores, have thus far been limited to cohort-level analyses, leaving its potential utility in providing individualised health estimates unexplored. Moreover, while MetaboHealth has been associated with incidence and prevalent morbidity in the UK Biobank[9], it remains unknown whether it also captures time to first chronic disease onset. Establishing this link is critical to assess whether predictors of mortality extend to morbidity, which together define the principal outcomes of ageing-related health. Finally, a molecular marker potentially supporting stratification of middle-aged individuals at risk of rapid overall health decline would ideally be actionable, i.e. showing a beneficial change in response to health-improving clinical or lifestyle interventions. Therefore, the purpose of this study is to address three main research questions: (1) Can MetaboHealth be transformed into a measure suitable for assessing the health of a single individual and be solely composed of clinically validated metabolic markers?; (2) Is MetaboHealth associated with age at first cardiometabolic disease onset?; (3) Does a lifestyle intervention change MetaboHealth beneficially to an extent that allows health economic profit calculations.

To address these questions, we used data from the Leiden Longevity Study (LLS), a three-generation (F0-2) cohort established in 2002[10]. We focused on the second generation (F2, *N* = 2,404) of LLS participants included, in which metabolomics data as well as long-term morbidity (up to 16 years) and mortality follow-up (up to 22 years) were collected. First, we investigated MetaboHealth score’s potential for individualized application by scaling the score based on an external reference distribution derived from the Biobanking and BioMolecular Research Infrastructure Netherlands (BBMRI-NL)[11]. In addition, we constructed a score using a clinically validated metabolite subset to test whether the predictive performance of MetaboHealth was retained. We used the updated mortality follow-up data through 2024 to determine the associations of the different MetaboHealth scores with all-cause mortality and examined whether the scores predicted age at first cardiometabolic disease onset. In addition, we examined the effects of our newly-generated Personal-MetaboHealth score on the age scale using Accelerated Failure Time (AFT) models. Finally, we investigated to what extent a short term (3-month) combined lifestyle intervention, i.e. the Growing Old Together (GOTO) study[12], changed the Personal-MetaboHealth score in individuals aged >60 years at risk of rapid overall health decline according to their baseline score.

## Methods

### Study Populations

#### Leiden Longevity Study

The LLS, initiated in 2002 in the Netherlands, is an ongoing cohort designed to investigate the molecular and environmental factors that may contribute to longevity. The study recruited long-lived siblings, men aged 89 years or older and women aged 91 years or older (F1), together with their offspring and the offspring’s partners (F2), resulting in a cohort of 421 long-lived families. We also collected demographic data of the parents of the F1 generation (i.e. F0). Here, we focused specifically on the F2 generation, that consists of 1,666 offspring and 738 partners, totaling 2,404 participants. Participants were followed for up max 22 years for mortality and max 16 years for morbidity. By 2018, 246 participants had died, ensuring that than 10.2% of the cohort was lost before morbidity follow-up ended. By 2025, 489 participants (20.3%) had died, while 498 (20.7%) had developed morbidity.

The cohort protocol was approved by the Medical Ethical Committee of the Leiden University Medical Center at 16th of August 2002, before the start of the study (P01.113), when the first participant enrolled on 5th of September 2002. In accordance with the Declaration of Helsinki, the LLS obtained informed consent from all participants prior to their entering the study.

#### Growing Old Together study

The GOTO study was a 13-week non-randomized, non-controlled, single-arm, open-label lifestyle intervention trial including 164 healthy older adults (83 males, 81 females, mean age 62.9 ± 5.7 years, mean BMI 26.9 ± 2.5 kg/m²). Participants underwent a 25% reduction in energy balance, achieved through a combination of a 12.5% decrease in caloric intake and a 12.5% increase in physical activity, as described by van de Rest and colleagues[12]. Prior to the intervention, baseline energy intake and expenditure were assessed using a 150-item food frequency questionnaire and the International Physical Activity Questionnaire–Short Form (IPAQ-SF). Personalized intervention guidelines were developed by a dietician and a physiotherapist in consultation with the participants, taking into account individual preferences and physical abilities. Dietary recommendations were designed to align as closely as possible with the Dutch Guidelines for a Healthy Diet. To promote adherence, participants were encouraged to increase physical activity in ways that fit their daily routines, with the aim of achieving full integration into regular daily life both during and after the intervention. Physical activities typically included walking, cycling, household tasks, and participation in local sports activities, either individually or with a partner. The primary outcome of the GOTO study was a significant reduction in fasting insulin levels, as previously reported[12].

The GOTO study protocol was approved by the Medical Ethical Committee of the Leiden University Medical Center before the start of the trial (P11.187). In accordance with the Declaration of Helsinki, the GOTO study obtained informed consent from all participants prior to their entering the trial. This trial was registered at the Dutch Trial Register (https://onderzoekmetmensen.nl/en/trial/27183) as GOT NL3301 and can also be found at the international clinical trials registry platform as NL-OMON27183. The first participant was enrolled on the 11^th^ of June 2012, while the last participant was enrolled on the 17^th^ of January 2013.

#### Biobanking and BioMolecular Research Infrastructure Netherlands

Another resource included in this project is the BBMRI-NL, a nationwide initiative that connects Dutch biobanks and cohort studies to support large-scale biomedical research. Established in 2009[11], it combines biological samples and data from a wide range of study populations: ALPHAOMEGA, BIOMARCS, CHARM, CHECK, DMS, DZS_WF, ERF, FUNCTGENOMICS, GARP, HELIUS, HOF, LIFELINES, LLS_PARTOFFS, LLS_SIBS, MRS, NESDA, PROSPER, RS, STABILITEIT, STEMI_GIPS-III, TOMAAT, UCORBIO, VUMC_ADC. Study information, including the ages at which biomaterial was obtained, is provided in our earlier work[13]and by the BBMRI consortium[11], including omics datasets such as metabolomics. All participating biobanks operate under ethical approval of their respective institutions, with informed consent obtained from all participants.

### Mortality Data

Since the beginning of the LLS, generation 3 mortality data has been updated through the Dutch Personal Records Database (BRP), a Dutch governmental service for identity information which is part of the nationwide Dutch population registry system (https://www.government.nl/topics/personal-data/personal-records-database-brp). This approach guarantees a consistent and reliable determination of vital status across the LLS-cohort.

### Morbidity Data

Morbidity data were available for a subset of LLS participants (N=1347; 56%) and were provided by the General Practitioners (GPs) of the third-generation participants. The morbidity data covers each participant’s disease history from birth until 2018, with GPs extracting information on age-related cardiometabolic diseases and the year of diagnosis directly from their health records. The records are routinely updated, even when individuals change doctors, ensuring continuity of information.

For this research, cardiometabolic diseases reported in the questionnaires were defined according to the International Statistical Classification of Diseases and Related Health Problems (ICD-10). The codes included were: transient ischemic attack (TIA; I60–I69), cerebrovascular accident (CVA; I63), angina pectoris (AP; I20), myocardial infarction (MI; I21), hypertension (I10), and diabetes (E10–E14).

### Questionnaire and pharmacy data

Information on smoking and drinking was obtained via a questionnaire administered during the entry phase of the study, while medication data were acquired directly from participants’ pharmacies. To calculate smoking quantity we multiplied the number of cigarettes per day by 365.25 and by the number of years smoked.

### Metabolomics and the MetaboHealth score

We used metabolomics data derived from EDTA plasma or serum samples collected at the time of recruitment of the study participants (LLS) or at the baseline and end of the intervention (GOTO). The samples were quantified by the Nightingale Health platform using ¹H-NMR spectroscopy. Using this method, Nightingale Health provided ∼250 circulating metabolites and derivates.

We focused on the 14 metabolites that together comprise the MetaboHealth score. This score was originally developed as a predictor of all-cause mortality and has since been shown to capture current health and frailty status[4]. The 14 metabolites composing the score include Acetoacetate (AcAce), Albumin (Alb), Glucose (Glc), Glycoprotein acetyls (Gp), Histidine (His), Isoleucine (Ile), Lactate (Lac), Leucine (Leu), Phenylalanine (Phe), the ratio of polyunsaturated fatty acids to total fatty acids (PUFA/FA), total lipids in small HDL (S-HDL-L), Valine (Val), mean diameter for VLDL particles (VLDL-D), and total lipids in chylomicrons and extremely large VLDL (XXL-VLDL-L). As the original MetaboHealth score was trained as an indicator of prospective mortality, higher levels indicate an increased mortality and health risk. Since we instead want to use MetaboHealth as a health indicator, we calculated its inverse so that increasing levels indicate increased survival and health.

### Statistical Analysis

All statistical analyses were performed with R (version 4.3.2), using the ‘FrailtyEM’ and ‘survival’ packages. A p-value threshold of <0.05 was considered statistically significant, and results are presented with corresponding 95% confidence intervals (CI).

We constructed three different MetaboHealth scores. First, the original MetaboHealth score was defined based on 14 metabolites: acetoacetate (AcAce), albumin (Alb), glucose (Glc), glycoprotein acetyls (Gp), histidine (His), isoleucine (Ile), lactate (Lac), leucine (Leu), phenylalanine (Phe), the ratio of polyunsaturated fatty acids to total fatty acids (PUFA/FA), total lipids in small HDL (S-HDL-L), valine (Val), mean diameter of VLDL particles (VLDL-D), and total lipids in chylomicrons and extremely large VLDL (XXL-VLDL-L); hereafter referred to as MetaboHealth. Second, using the same 14 metabolites, we developed an individualized MetaboHealth score based on BBMRI-NL data, hereafter referred to as Rescaled-MetaboHealth. Finally, we constructed a clinically validated individualized MetaboHealth score based on 10 clinically validated metabolites (PUFA/FA, Glc, Alb, Gp, Phe, Ile, Leu, Val, His, and Lac), hereafter referred to as Personal-MetaboHealth.

The script used to calculate the MetaboHealth scores from raw Nightingale metabolite data is provided here as free, open-access material: https://git.lumc.nl/publications/individualized-metabohealth.

In a subsequent step, we focused on the F2 generation of the LLS data and z-scaled the three MetaboHealth scores so that they have a mean of 0 and a standard deviation of 1. This standardization enabled direct comparison of effect sizes across different analyses. As a first analytical step, we used Cox random-effects (frailty) survival models to examine the extent to which the scores were associated with prospective mortality and morbidity and to identify which score variant showed the strongest effect. The analyses in step one were adjusted for sex and age at inclusion, with age at inclusion included as a left-truncation point in the survival analyses. In step two, we repeated the analyses from step one with additional adjustment for smoking quantity, alcohol consumption, and medication use.

To estimate how the different the Personal-MetaboHealth score associate with time to morbidity on the age scale, we used Accelerated failure time (AFT) models. Unlike Cox proportional hazards models, AFT models directly quantify how covariates accelerate or decelerate the expected time until an event. This allows results to be interpreted in terms of survival time. In our analyses, survival time was defined as the difference between age at study entry and age at first disease or last follow-up. The AFT models were adjustment for sex, age at study entry, and familial clustering. Several parametric distributions were tested (exponential, Weibull, Gaussian, logistic, log-logistic, and log-normal) and model fit was evaluated using Akaike (AIC) and Bayesian (BIC) information criteria. The Exponential distribution provided the best fit for mortality.

Finally, we used the GOTO study to investigate whether, and to what extent, the PersonalMetaboHealth score improves following the intervention. Participants were older adults who underwent a 13-week lifestyle program combining reduced caloric intake and increased physical activity, with personalized guidance from a dietician and physiotherapist. We analyzed the changes in Personal-MetaboHealth using paired t-tests, comparing baseline and post-intervention scores to assess the impact of the lifestyle changes on overall health.

## Results

Our initial analyses were conducted within the LLS cohort, which includes data on three generations: long-lived siblings (F1), their deceased parents (F0), and their middle-aged offspring with partners (F2), where the F1 and F2 generations are part of the LLS Biobank. For the present study, we focused on the F2 generation, consisting of 1,666 offspring and 738 partners, leading to a grand total of 2,404 participants. At baseline, participants had a mean age of 59.2 years and were followed for up to 22 years for all-cause mortality and 16 years for cardiometabolic disease incidence (**Table 1**).

**Table 1.** Demographic characteristics from the F2 participants of the LLS.

| Characteristics |  |
| --- | --- |
| Participants, *N* | 2,404 |
| Offspring/partners, *N* | 1,666/738 |
| Age at inclusion, mean (SD) | 59.1 (6.8) |
| Female, *N* (%) | 1,326 (55.2) |
| Follow-up time mortality, mean (SD) | 18.1 (3.9) |
| Deceased, *N* (%) | 489 (20.3) |
| Age at death, mean (SD) | 76.3 (8.6) |
| Disease free at inclusion, *N* (%) | 1,403 (58.4) |
| Follow-up time cardiometabolic disease, mean (SD) | 10.1 (3.7) |
| Cardiometabolic disease, *N* (%) | 433 (30.9) |
| Metabolomics data available, *N* (%) | 2,301 (95.7) |
| Smoking, *N* (%) | 1,316 (54.7) |
| Smoking quantity (per 10 cigarettes/day), mean (SD) | 8,015.6 (11,957.1) |
| Alcohol consumption, mean (SD) | 14.6 (14.8) |
| Medication use, *N* (%) | 507 (21.1) |

### Creation of individualised MetaboHealth scores

A key aim of this study was to evaluate whether our original MetaboHealth score could be adapted to estimate health status at the individual level. To this end, we compared three versions of the score, including the original one. To enable application at the individual level, we rescaled the MetaboHealth score using metabolite reference distributions from the nationwide BBMRI-NL resource[11], which is a national node of the European BBMRI consortium (BBMRI-ERIC), representing a geographic distribution of the Dutch population, with a strong, although not exclusive, Caucasian focus. We termed this score Rescaled-MetaboHealth. Next, we aimed to enhance clinical applicability by restricting the Rescaled-MetaboHealth to the 10 metabolites that are clinically validated by Nightingale Health (see methods), thereby excluding cholesterol-related metabolites and acetoacetate and termed this score Personal-MetaboHealth. To improve interpretability, we rescaled all MetaboHealth scores in a way that a higher score indicates better health.

### The different MetaboHealth scores associate equally well with all-cause mortality

As a first step, we compared the association of the different versions of MetaboHealth score with prospective mortality. Our results indicate replication of our previous work, as we observed a strong association of the original MetaboHealth score with all-cause mortality in the LLS[4]. We showed that each standard deviation increase in the score corresponded to a 23% lower risk of death (HR = 0.77, 95% CI: 0.71–0.83, P = 4.58 × 10□^1^□, **Figure 1**). The mortality associations of the Rescaled-MetaboHealth (HR = 0.76, 95% CI: 0.70–0.83, P = 2.24 × 10□^1^□, **Figure 1**) and Personal-MetaboHealth scores (HR = 0.75, 95% CI: 0.69–0.82, P = 5.31 × 10□^1^□, **Figure 1**) were of similar strength, indicating that the predictive performance was preserved when the score was standardised to external data and confined to the clinically validated subset of the original variables.

**Figure 1.**
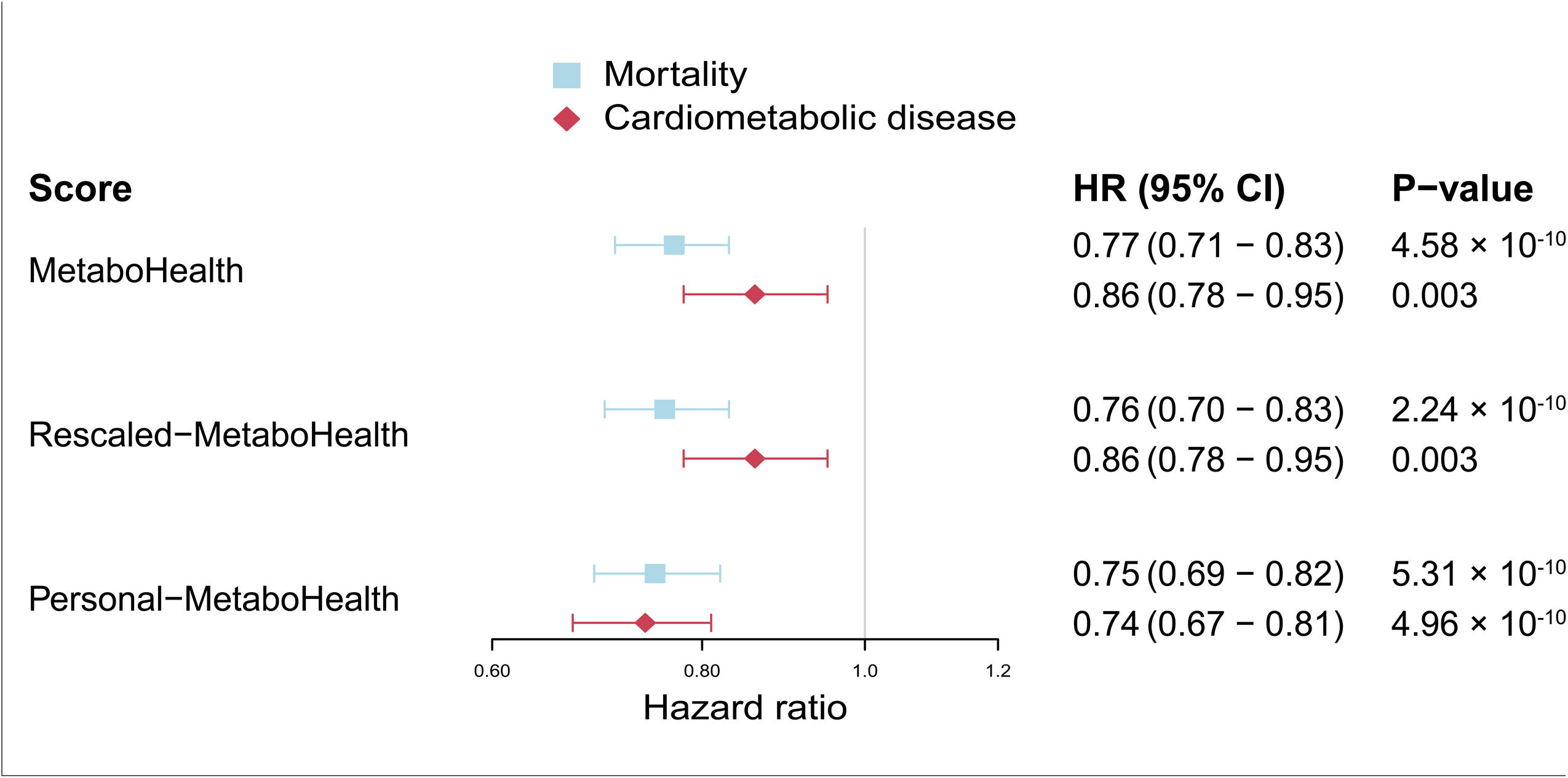
Association between the different MetaboHealth scores and all-cause mortality and cardiometabolic disease morbidity. The figure depicts the different versions of MetaboHealth in association with time to mortality (squares) and time to first cardiometabolic disease (diamond). The statistical models are adjusted for age, sex, and family relations using a gaussian random effect. HR=hazard ratio; CI=confidence interval.

### Personal-MetaboHealth is most strongly associated with cardiometabolic disease

As a second step, we compared the association of the MetaboHealth scores with the onset of first cardiometabolic disease. To this end, we used the 1,403 participants from the LLS who were free of cardiometabolic disease at baseline. We observed that higher levels of all three MetaboHealth scores were strongly associated with a delayed onset of first cardiometabolic disease (**Figure 1**). Notably, the strongest effect was observed for Personal-MetaboHealth, which showed a 26% decreased yearly risk of developing a first cardiometabolic disease with each increase in standard deviation (HR = 0.74, 95% CI: 0.67–0.81, P = 4.96 × 10□^1^□). The effect of the Personal-MetaboHealth score was 12% stronger (HR = 0.74 vs. HR = 0.86) compared to the other two MetaboHealth scores.

### Adjustment for covariates attenuates the effects of MetaboHealth

Given that previous studies have shown a strong association of the original MetaboHealth score with lifestyle factors[14–16], we performed robustness checks in which we adjusted for smoking quantity, alcohol consumption, and medication use. For all MetaboHealth scores the strength of the association with all-cause mortality was slightly attenuated, but remained statistically significant (P ≤ 0.006, **Table 2**). For cardiometabolic disease incidence, however, the associations of both the original MetaboHealth and Rescaled-MetaboHealth were no longer significant (P = 0.079 and P = 0.073, respectively), while the association of Personal-MetaboHealth was also attenuated (HR_Adjusted_ = 0.78 vs HR_Unadjusted_ = 0.74), but remained significant (P = 1.92 × 10^-4^, **Table 2**). Sensitivity analyses showed that the majority of the reduction in the association between the MetaboHealth scores and all-cause mortality and cardiometabolic disease incidence is caused by adjustment for smoking quantity (**Supplementary Table 1**). This indicates that the scores partly cover the negative effects of smoking on health.

**Table 2.** MetaboHealth scores, all-cause mortality, and cardiometabolic disease incidence.

|  | Basic (*N* = 2,301) |  | Adjusted (*N* = 1,228) |  |
| --- | --- | --- | --- | --- |
|  | HR (95% CI) | P-value | HR (95% CI) | P-value |
| **Mortality** |  |  |  |  |
| MetaboHealth | 0.77 (0.71 - 0.83) | $4.58 \times 10^{-10}$ | 0.81 (0.70 - 0.93) | 0.004 |
| Rescaled-MetaboHealth | 0.76 (0.70 - 0.83) | $2.24 \times 10^{-10}$ | 0.79 (0.69 - 0.92) | 0.002 |
| Personal-MetaboHealth | 0.75 (0.69 - 0.82) | $5.31 \times 10^{-10}$ | 0.82 (0.71 - 0.94) | 0.006 |
| **Cardiometabolic disease** |  |  |  |  |
| MetaboHealth | 0.86 (0.78 - 0.95) | 0.003 | 0.89 (0.78 - 1.01) | 0.079 |
| Rescaled-MetaboHealth | 0.86 (0.78 - 0.95) | 0.003 | 0.89 (0.78 - 1.02) | 0.073 |
| Personal-MetaboHealth | 0.74 (0.67 - 0.81) | $4.96 \times 10^{-10}$ | 0.78 (0.68 - 0.89) | $1.92 \times 10^{-4}$ |

HR, hazard ratio; 95% CI, 95% confidence interval. The basic models are adjusted for age, sex, and family relations (random effect), while the adjusted models are additionally adjusted for smoking, alcohol consumption, and medication use.

|  | Basic (*N* = 2,301) |  | Adjusted (*N* = 1,228) |  |
| --- | --- | --- | --- | --- |
|  | HR (95% CI) | P-value | HR (95% CI) | P-value |
| **Mortality** |  |  |  |  |
| MetaboHealth | 0.77 (0.71 - 0.83) | 4.58 x 10<sup>-10</sup> | 0.81 (0.70 - 0.93) | 0.004 |
| Rescaled-MetaboHealth | 0.76 (0.70 - 0.83) | 2.24 x 10<sup>-10</sup> | 0.79 (0.69 - 0.92) | 0.002 |
| Personal-MetaboHealth | 0.75 (0.69 - 0.82) | 5.31 x 10<sup>-10</sup> | 0.82 (0.71 - 0.94) | 0.006 |
| **Cardiometabolic disease** |  |  |  |  |
| MetaboHealth | 0.86 (0.78 - 0.95) | 0.003 | 0.89 (0.78 - 1.01) | 0.079 |
| Rescaled-MetaboHealth | 0.86 (0.78 - 0.95) | 0.003 | 0.89 (0.78 - 1.02) | 0.073 |
| Personal-MetaboHealth | 0.74 (0.67 - 0.81) | 4.96 x 10<sup>-10</sup> | 0.78 (0.68 - 0.89) | 1.92 x 10<sup>-4</sup> |

HR, hazard ratio; 95% CI, 95% confidence interval. The basic models are adjusted for age, sex, and family relations (random effect), while the adjusted models are additionally adjusted for smoking, alcohol consumption, and medication use.

### One standard deviation increase in Personal-MetaboHealth delays cardiometabolic disease onset by over a decade

Instead of quantifying the effect of a marker on an outcome as a hazard ratio, it is also possible to quantify this at the age scale, i.e. the number of years of later/earlier onset of an outcome, using AFT models, which allows more intuitive interpretation. When applied to Personal-MetaboHealth, the AFT model showed a ‘time ratio’ of 1.34 (95% CI: 1.23–1.47; P = 1.73 × 10□^1^□) which indicates that with every SD increase in Personal-MetaboHealth the onset of first cardiometabolic disease is decelerated by 34%. This translates to an 11.7 year later onset of the first cardiometabolic disease per increase in SD of the score considering an average person within our LLS dataset.

### Personal-MetaboHealth positively responds to a lifestyle intervention

For implementation of a biomarker as a health check in population health, it needs to be actionable, i.e. modifiable through an evidence-based intervention. We therefore investigated to what extent a 3-month combined lifestyle intervention (GOTO study) changed the Personal-MetaboHealth score in middle aged individuals (59.1 years (SD 6.8). When we combined the data from all individuals within the study, we observed no change in the Personal-MetaboHealth score due to the intervention (β = 0.01, 95% CI: −0.03–0.06, P-value = 0.666). We previously observed that the GOTO intervention in the overall quite healthy middle-aged participants, is most effective in improving health-related variables, such as systolic blood pressure, in individuals that were the most unhealthy at baseline[12]. We therefore stratified the GOTO participants into two groups (<0, ‘unhealthy’ (n = 52) and ≥0, ‘healthy’ (n = 83)) based on the baseline Personal-MetaboHealth scores. We observed that the individuals in the unhealthy group significantly improved their score as a results of the intervention (β = 0.15, 95% CI: 0.05–0.24, P-value = 0.003), while the opposite was observed, although to a lesser extent, for the individuals in the healthy group (β = −0.07, 95% CI: −0.12–-0.02, P-value = 0.007). Moreover, we observed a difference between the two groups in other health-related parameters in response to the intervention, such as fasting triglyceride, CRP, and insulin levels (the primary outcome of the GOTO study). On the other hand, markers of body composition and blood pressure showed a similar change in both groups (**Figure 2**). This indicates that the Personal-MetaboHealth score is an actionable biomarker that can be altered through an effective lifestyle intervention in middle-aged at risk individuals.

**Figure 2.**
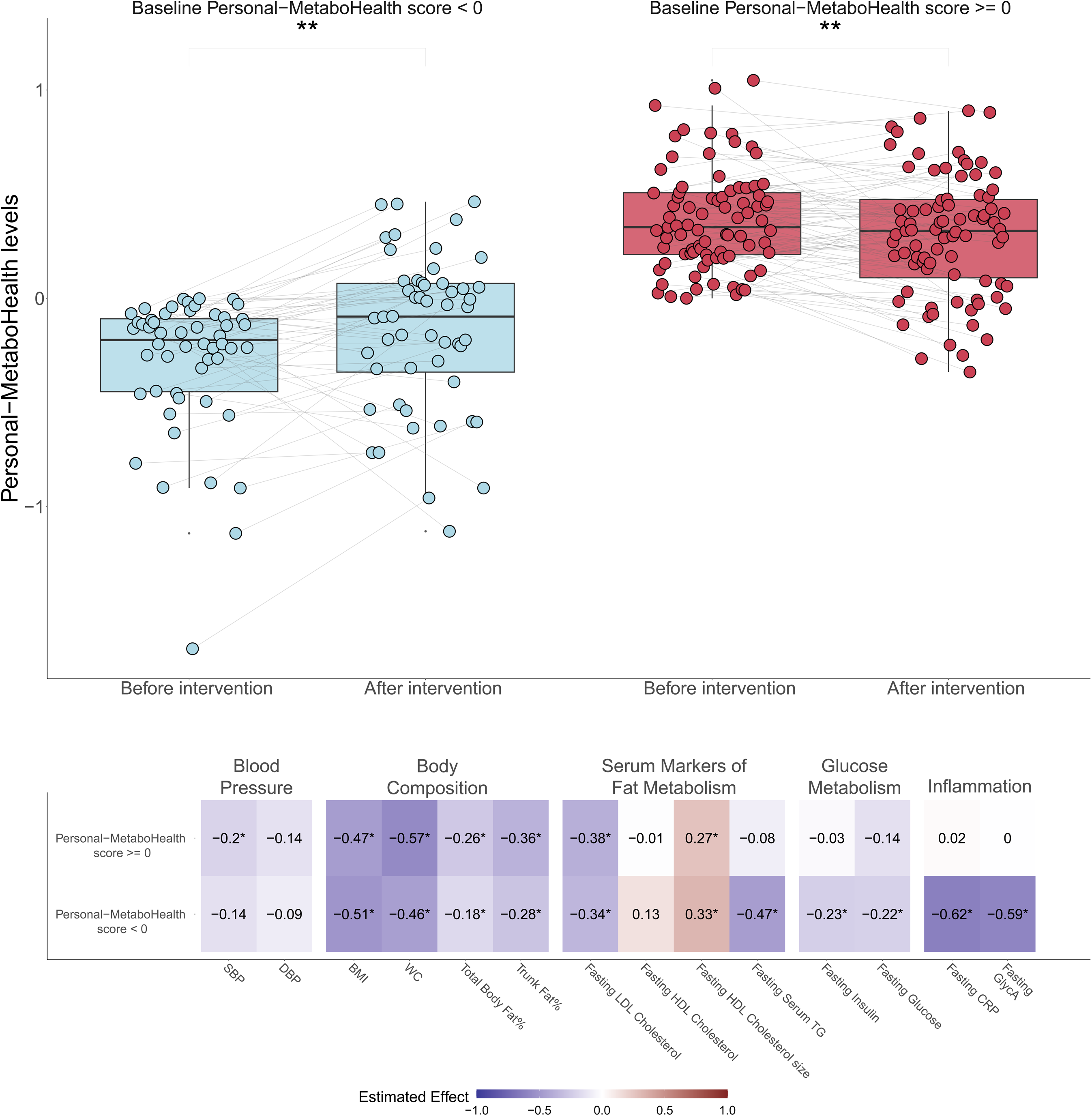
Changes in Personal-MetaboHealth scores, stratified by baseline scores. The figure depicts the change in Personal MetaboHealth in GOTO for those with a baseline MetaboHealth below zero (top left) and for those with a PersonalMetaboHealth above or equal to zero (top right). Analyses were done using a mixed model with a gaussian random effect to model individual changes over time. The analyses are adjusted for age and sex at baseline as fixed effects (before intervention) and family-id and person-id as random effects. The significance level is represented by an asterisk: * p-value < 0.05, ** p-value < 0.01. The bottom shows the association between the z-scaled health markers and the intervention time point for participants stratified at baseline (above and below Personal MetaboHealth of 0). These analyses are also adjusted for age and sex at baseline as fixed effects (before intervention) and family-id and person-id as random effects. The values depicted represent the estimated effect between the z-scaled health marker and intervention time point, this is also represented by the colors: blue represents a negative effect, red represents a positive effect, white represents a weak effect. An asterisk indicates that the association is significant after Benjamini-Hochberg correction for multiple testing.

## Discussion

In this study, we investigated whether the MetaboHealth score, a biomarker of overall health, could be modified so it can be used as an actionable health check in middle age for prevention strategies. We created different versions of MetaboHealth, one that is individualized by scaling based on the Dutch population (Rescaled-MetaboHealth) and one that is additionally confined to the subset of clinically validated metabolic biomarkers (Personal-MetaboHealth). We subsequently showed that all three scores perform equally well in predicting prospective all-cause mortality. However, Personal-MetaboHealth outperforms the other two scores in predicting time to first cardiometabolic disease and does so independently of smoking, alcohol consumption and medication use. A one standard deviation increase in the score is associated with a 11.7 year later onset of the first cardiometabolic disease. Moreover, we showed that Personal-MetaboHealth can be improved by a combined lifestyle intervention (GOTO study), specifically in at risk individuals with unhealthy baseline scores. Together, these results establish Personal-MetaboHealth as a promising health check for individualised risk stratification and response to lifestyle interventions and therefore as a potential tool to guide preventive strategies in population health.

The original MetaboHealth score was created using metabolic markers that were z-scaled separately per study population[4]. This was needed because the 12 used studies could not be analysed together, while we wanted to perform a direct comparison between metabolic markers measured on different scales and ranges, facilitating feature selection based on effect size, and ensure that no single feature dominated the construction of the score. However, this means original MetaboHealth scores cannot be compared between individuals from different populations, limiting its usability. To overcome this problem, we scaled the metabolic markers using a Dutch reference population (BBMRI-NL) consisting of 29,792 participants from 24 cohorts with age ranges from 16 to 103 years. This individualisation of the MetaboHealth score now allows direct comparisons of effects on health-related outcomes between individuals from different populations.

Over the last couple of years, Nightingale Health has clinically validated over 50 of their metabolic markers. The advantage of restricting the Personal-MetaboHealth score to the 10 clinically validated metabolic markers that were part of the original MetaboHealth score is that this would make the score more attractive for use in population health and clinical settings, because it would meet all regulatory requirements. Interestingly, the Personal-MetaboHealth outperformed the original MetaboHealth score in predicting cardiometabolic disease incidence, which we hypothesise to be partly due to the removal of metabolic biomarkers with more technical variability, such as acetoacetate and S-HDL-L[17]. Future studies will have to show if this is something specific for cardiometabolic disease or if the same applies to other health-related outcomes.

We quantified the effects of Personal-MetaboHealth as a change in hazard as well as on the age scale (using an AFT model). The advantage of the latter is that it provides a more intuitive interpretation that can be coupled back to an individual. These analyses showed a substantial and similar effect of Personal-MetaboHealth on both all-cause mortality (33% change in risk per change in standard deviation) and cardiometabolic disease onset (35% change in risk, or 11.7 year difference in onset, per change in standard deviation), of which the magnitude is comparable to well-known risk factors such as socioeconomic status and smoking[18,19]. Interestingly, the association between the original MetaboHealth score and cardiometabolic disease incidence seems to be largely explained by the effects of smoking, which is not surprising given the previously reported effects of smoking on both MetaboHealth[14] and, especially, cardiovascular disease[20]. However, the association between Personal-MetaboHealth and cardiometabolic disease incidence is largely independent of smoking, indicating that this new score captures broader aspects of overall health than those affected by smoking.

Evidence regarding the effectiveness of lifestyle interventions in older adult populations remains inconclusive. Therefore, we previously performed the GOTO study and showed that this combined diet and exercise intervention is able to improve overall health in individuals aged between 46 and 75 years[20]. However, we observed a large heterogeneity in the response of individuals to the intervention, which was partly explained by difference in physical behaviour during the intervention[21]. In the current study, we found that another part of the heterogeneity in response may be due to differences in Personal-MetaboHealth scores at baseline. We showed that individuals with unhealthy Personal-MetaboHealth scores were able to improve their score during the intervention, which was coupled to improvements in other health parameters, such as blood pressure and markers of cholesterol metabolism. This indicates that the Personal-MetaboHealth score may be particularly useful for identification and monitoring of at risk middle aged adults in need of a lifestyle intervention.

A strong focus in the field of biomarkers of ageing has been on CpG methylation-based scores, of which the mortality predictor GrimAge showed the most robust effects within the GOTO study[22]. Although the intervention effect on Personal-MetaboHealth only becomes apparent after stratifying individuals based on their baseline values, GrimAge declines in all individuals. While the decline in GrimAge is most strongly associated with a change in parameters related to body composition, we observed that the changes in individuals with unhealthy Personal-MetaboHealth scores seems to represent broader immune-metabolic changes, especially in fat metabolism and inflammation[22]. This indicates that metabolomics- and CpG methylation-based mortality predictors seem to capture different aspects of intervention-based health improvements and should ideally be used in combination.

A key strength of our study is the use of the prospective design of the LLS, with follow up for all-cause mortality of up to 22 years and cardiometabolic disease onset of up to 16 years. Detailed information on disease incidence and medication use was obtained directly from participants’ pharmacies and general practitioners, ensuring reliable outcome ascertainment. However, a limitation of the LLS is that the study design is complex due to its focus on families and the presence of right censoring and left truncation. Currently, no statistical method adequately accounts for both familial correlation and left truncation when estimating AFT models. This may impact the baseline hazards of the AFT models and thereby complicate interpretation on the age scale.. Additionally, metabolomic data was only assessed at baseline, preventing the modelling of temporal changes. These limitations narrow the scope of inference but also highlight priorities for future research. Replication in larger, more diverse cohorts and the inclusion of longitudinal metabolomic trajectories will be essential to validate and extend these findings, but this was out of scope for the current study. Another limitation is that the scaling of Rescaled-MetaboHealth and Personal-MetaboHealth was based on BBMRI-NL, which is a reference population that mainly consists of Caucasian individuals and may thus provide an incomplete representation of the metabolomic diversity in other countries and ethnic populations, potentially, even within the Netherlands. As a next step we therefore plan to test the validity of the Personal-MetaboHealth in populations that are currently underrepresented within BBMRI. Moreover, future studies can use a similar approach to create country-specific Personal-MetaboHealth variations based on large reference populations with Nightingale Health data.

In summary, we created an individualised version of the MetaboHealth score, which we termed Personal-MetaboHealth. In comparison to the original score, Personal-MetaboHealth retains its robust association with all-cause mortality in the LLS, while it shows an even stronger association with cardiometabolic disease onset, which was independent from smoking, alcohol consumption and medication use. Moreover, using data from the GOTO study, we found that Personal-MetaboHealth can be used to identify and monitor at risk middle aged individuals in need of a lifestyle intervention. Replication in larger and more diverse cohorts will be essential to confirm these associations and to establish applicability of Personal-MetaboHealth in different healthcare settings. Ultimately, integrating the Personal-MetaboHealth score into population health frameworks as an actionable health check in middle age may offer a powerful approach for early disease prevention and extension of healthy lifespan.

## Competing interest

The authors declare no competing interests.

## Acknowledgments

The construction and maintenance of the LLS data has received funding from the European Union’s Seventh Framework Programme (FP7/2007-2011) under grant agreement number 259679. This study was further supported by the Netherlands Consortium for Healthy Ageing (grant 050-060-810), in the framework of the Netherlands Genomics Initiative, Netherlands Organization for Scientific Research (NWO)BBMRI-NL, a Research Infrastructure financed by the Dutch government (NWO 184.021.007 and 184.033.111). Niels van den Berg obtained funding through the research project “An Age Old Advantage?” (P21-0139), the Swedish Foundation for the Humanities and Social Sciences (Riksbankens Jubileumfond, RJ) and the Netherlands Organization for Scientific Research, domain Health Research and Medical Sciences (09120012010052).

## Author contributions

Niels van den Berg, Joris Deelen, and P. Eline Slagboom conceived the project. Gabriela Natalle Lopes conducted the majority of the hands-on research as part of her master’s internship, under the supervision of Joris Deelen and Niels van den Berg. Edson Amaro Junior provided internal supervision from Brazil and supported Gabriela Natalle Lopes during her research stay in the Netherlands. Fatih Bogaards conducted the GoTo analysis. Niels van den Berg and Joris Deelen finalized the analyses, performed additional hands-on work, and jointly prepared the first draft of the manuscript based on Gabriela Natalle Lopes’ internship report. All authors contributed to and approved the final version of the manuscript for publication.

CRediT taxonomy–based Author Contributions Author Contributions (CRediT):

- Conceptualization: Niels van den Berg, Joris Deelen, P. Eline Slagboom
- Supervision: Joris Deelen, Niels van den Berg, Edson Amaro Junior
- Investigation: Gabriela Natalle Lopes
- Formal Analysis: Niels van den Berg, Joris Deelen, Fatih Bogaards
- Methodology: Niels van den Berg and Joris Deelen
- Writing – Original Draft: Niels van den Berg, Joris Deelen
- Writing – Review & Editing: Niels van den Berg, Gabriela Natalle Lopes, Fatih Bogaards, Marian Beekman, Edson Amaro Junior, Joris Deelen, P. Eline Slagboom

## Code availability

The scripts containing the code for data pre-processing, data analyses, and MetaboHealth construction can be freely downloaded at: https://git.lumc.nl/publications/individualized-metabohealth.

## Data availability

The individual-level data from the LLS are protected by Dutch personal integrity laws, and other (privacy) regulations. As such, restrictions apply to the availability of the LLS data, which were used under license for the current study, and so are not publicly available. For both datasets, summary statistics are available upon request to the corresponding authors (Niels van den Berg and Joris Deelen). The LLS data is available following a data access procedure (https://leidenlangleven.nl/data-access/). Each request will be evaluated whether the research is compliant with the informed consent that has been signed by the LLS participants.

## Notes

### Competing Interest Statement

The authors have declared no competing interest.

### Clinical Trial

NL-OMON27183

### Funding Statement

The construction and maintenance of the LLS data has received funding from the European Union Seventh Framework Programme (FP7/2007-2011) under grant agreement number 259679. This study was further supported by the Netherlands Consortium for Healthy Age-ing (grant 050-060-810), in the framework of the Netherlands Genomics Initiative, Nether-lands Organization for Scientific Research (NWO)BBMRI-NL, a Research Infrastructure financed by the Dutch government (NWO 184.021.007 and 184.033.111). Niels van den Berg obtained funding through the research project An Age Old Advantage? (P21-0139), the Swedish Foundation for the Humanities and Social Sciences (Riksbankens Jubile-umfond, RJ) and the Netherlands Organization for Scientific Research, domain Health Re-search and Medical Sciences (09120012010052).

### Author Declarations

The Leiden Longevity Study cohort protocol was approved by the Medical Ethical Committee of the Leiden Univer-sity Medical Center at 16th of August 2002, before the start of the study (P01.113), when the first participant enrolled on 5th of September 2002. In accordance with the Declaration of Helsinki, the LLS obtained informed consent from all participants prior to their entering the study. The GOTO study protocol was approved by the Medical Ethical Committee of the Leiden University Medical Center before the start of the trial (P11.187). In accordance with the Declaration of Helsinki, the GOTO study obtained informed consent from all participants prior to their entering the trial. This trial was registered at the Dutch Trial Register (https://onderzoekmetmensen.nl/en/trial/27183) as GOT NL3301 and can also be found at the international clinical trials registry platform as NL-OMON27183.

### Summary of Updates

Update of a small error in the metabolites composing the personal metabohealth and the affiliation of prof Edson Amaro Junior.

